# Neurophysiological dysconnectivity across multiple resting state brain networks and cognitive impairment in children with Prader-Willi Syndrome

**DOI:** 10.1101/2025.02.25.25322869

**Authors:** Benjamin T Dunkley, Kevin G Solar, Rouzbeh Zamyadi, Amy C Reichelt, Ellie Morrison, Shannon E Scratch, Jill Hamilton

## Abstract

Prader-Willi Syndrome (PWS) is a rare genetic condition with multifaceted physical, behavioural and cognitive difficulties that is characterized by hyperphagia and low executive functioning. Food-seeking behaviours may be moderated by hormonal, cognitive, and psychological factors, and are thought to be mediated in part by functional brain abnormalities. Here, we used an experimental protocol integrating eyes opens resting state magnetoencephalography (MEG) - a high-resolution neurophysiological imaging technique - and neuropsychological profiling to understand the relationship between executive functioning, and intrinsic brain activity & functional connectivity in a prospective, cross-sectional cohort with PWS, and a sex-, age- and BMI-matched control group. We observed lower executive functioning in PWS as well as functional dysconnectivity across multiple channels of brain synchrony - in other words, across multiple frequency bands that mediate communication within and between brain networks - in the visual, attentional, and the default mode networks. Moreover, we found ‘brain-wide’ changes in the topological structure of brain networks in those with PWS, with increased ‘hubness’ of functional networks, but decreased centrality. However, none of these measures survived multiple comparison correction after correlating with neuropsychological outcomes, although there were moderate effect sizes (degree of association). This is the first study to combine neuropsychology and neurophysiological imaging to show that functional synchrony in multiple brain networks is dysregulated in PWS.

## Introduction

Prader-Willi Syndrome (PWS) is a rare genetic disorder, most commonly due to a paternal deletion of a region on chromosome 15 (DEL) or maternal uniparental disomy (UPD). It is characterised by behavioural, cognitive, endocrine, and physical challenges. The phenotype of PWS progresses throughout development. Infants initially present with hypotonia, poor feeding ability, and failure to thrive (Cassidy et al., 2012; Driscoll et al., 1993). By early childhood, hyperphagia and food-seeking behaviour emerge, increasing the risk for severe obesity and associated complications if access to food is left uncontrolled (Manning & Holland, 2015).

Global developmental delay is present, and physical and social milestones are reached around twice the normal age (Costa et al., 2019). Many individuals with PWS show mild to moderate intellectual disability, lower intelligent quotient (IQ), and impairments in language, social relationships, academic performance, and executive functioning (Costa et al., 2019; Qiu et al., 2024; Skokauskas et al., 2012; Whittington & Holland, 2017). Psychiatric symptoms such as obsessive-compulsive tendencies, anxiety, and temper outbursts are also common (Schwartz et al., 2021). Growing evidence suggests that these characteristic symptoms of PWS are driven in part by functional and structural brain abnormalities.

The endocrine and metabolic dysfunction in PWS is indicative of abnormalities in the hypothalamus (Burman et al., 2001; Eiholzer et al., 2006). To date, food-seeking and hyperphagia has been a focus of neuroscientific research in PWS. Compared to controls, children and adolescents with PWS (9-19 years old) display dysregulated functioning measured with functional MRI (fMRI) in reward circuits, with regions including the amygdala, insula, and nucleus accumbens (Holsen et al., 2006). Children with PWS also exhibit altered resting-state functional connectivity using fMRI in appetite and reward networks, such as the prefrontal cortex (PFC) and between the hypothalamus and lateral occipital complex (LOC) (Lukoshe et al., 2017; Zhang et al., 2013). The latter may partly explain the preoccupation with food seen in PWS, given the LOC’s role in processing visual food cues (Lukoshe et al., 2017).

The neural underpinnings of the cognitive profile in PWS are less understood; however, its manifestations are suggestive of atypical brain development beyond the hypothalamus (Manning & Holland, 2015). Most research to date has related cognition to brain structure rather than function in PWS. Reduced white and grey matter volume in several brain regions has been postulated to partially account for IQ, attentional, and executive function deficits seen in this group (Whittington & Holland, 2017). Moreover, lower cortical complexity, evidenced by aberrant cortical gyrification and its correlation with IQ in children with PWS (6-19 years old), may play a role in impaired cognition and development in PWS (Lukoshe et al., 2014). Recent structural-functional data suggests that individuals with PWS present with decreased structural-functional correlations in brain networks and altered topological architecture that correlate with developmental outcomes (Huang et al., 2024).

Understanding how these cognitive challenges map onto functional brain systems remains relatively unknown. One electroencephalogram (EEG) study found that adults with PWS had diminished P3 wave activity, a metric of attention, memory, decision-making and cognitive function, in auditory and visual pathways compared to controls (Borra & Magosso, 2021; Stauder et al., 2002). Another study using magnetoencephalography (MEG) examined somatomotor evoked responses in PWS, and found evidence of deficits in gamma-aminobutyric acid type A receptor subunit (GABAaR) genes (Egawa et al., 2021). Additionally, atypical functional activation in the parietal and prefrontal cortices, using fMRI, may contribute to task-switching difficulty seen in PWS (Whittington & Holland, 2017). Altered functional connectivity strength in the default mode, motor-sensory, and prefrontal networks have been implicated in eating and reward in people with PWS (Zhang et al., 2013), however whether altered functional connectivity underpinning cognitive and executive functioning has yet to be explored in PWS.

Examining brain connectivity in systems involved in food reward and cognitive control and its relationship to neuropsychological performance provides a foundation for the development of effective treatment therapies to manage eating behaviour and chronic stress in PWS. Brain imaging in the resting state is an effective methodology for participants with cognitive impairments, as it does not require comprehension of complex task instructions (Lukoshe et al., 2017). Similarly, using parent-report measures of behaviour and executive function may be more accurate than self-report measures in this population.

Here, we use MEG - a non-invasive, functional brain imaging technique, that is well-tolerated by children with behavioural challenges - to measure intrinsic brain functioning to advance upon previous investigations. MEG directly measures neural activation, using a millisecond timescale, allowing us to reveal fast activation dynamics and understand the neurophysiological mechanisms that relate to executive dysfunction in this group. MEG is particularly strong at imaging functional connectivity and allows us to critically relate brain activity and circuitry to cognitive outcomes in PWS. We are the first group to use this technology to understand functional connectivity in the PWS population. We also recruited BMI-matched controls as studies have shown that BMI is associated with significant changes in neurophysiological functioning in youth (Reichelt et al., 2024). We hypothesised that adolescents with PWS will exhibit reduced functional connectivity compared to BMI-matched controls amongst frontal and fronto-striatal regions, particularly areas that form part of the default motor and central executive networks, including the dorsolateral and dorsomedial prefrontal areas, and that reduced connectivity amongst frontal and fronto-striatal regions are related to deficits in executive functioning.

## Methods

### Participants

Participant data collection occurred from 2019 to 2022, and along with all analyses, was conducted at the Hospital for Sick Children in Toronto, ON, Canada. Participants were recruited from the Healthy Living and Endocrinology clinics at the Hospital for Sick Children in Toronto and from the community, as part of a prospective cross-sectional cohort study. Initially, n=35 total participants were recruited; two participants were removed due to excessive head motion resulting in very few trials remaining. MEG data for two additional participants were not collected due to scanner malfunction. For all subsequently reported MEG and outcome analyses: n=15 PWS group with a genetically confirmed diagnosis (8 DEL, 4 UPD, 3 unknown; aged 7 to 16 years; mean 12.6 ± 2.6 years; 6 females; mean BMI z-score 2.43) and n=16 BMI-matched control group (aged 8 to 16 years; mean 12.8 ± 2.6 years; 7 females; mean BMI z-score 2.53). The groups were not significantly different in age (p =0.08), nor sex ratios (Chi-Square test, p = 0.83), nor z-scored BMI (Kruskal-Wallis test, p = 0.784).

Participants were eligible for this study if they were between 7-16 years of age, had sufficient English language skills and cognitive capacity to understand task instructions, and had no contraindications to MRI or MEG scanning. This study was approved by the Hospital for Sick Children Ethics Board. Written informed consent was obtained from all caregivers of participants in accordance with the Declaration of Helsinki. Participants assented when required by the Hospital for Sick Children Ethics Board.

All data collection occurred during two separate study visits at the Hospital for Sick Children that were performed about two weeks apart on average. At study visit 1, height and weight were measured while fasting, without shoes, and in light clothing using a digital scale and stadiometer. Sex and age specific BMI percentiles and z-scores were determined using the WHO growth chart for Canadian children (*WHO Growth Charts for Canada*, 2019). After a standardised breakfast, participants completed the MEG neuroimaging session (1.5 hours). At study visit 2, participants completed the neuropsychological assessments (3 hours total) with breaks offered every 30 minutes to avoid fatigue. Participants were asked to eat and feel as full as they would normally for both study visits.

### Neurobehavioural and psychiatric outcomes

Children in the PWS and control groups completed core subtests from the Wechsler Intelligence Scale for Children, Fifth Edition (WISC-V) (Cormier et al., 2016), and their parents completed the parent-forms of the Behavior Assessment System for Children, Third Edition (BASC-3) (Thorpe et al., 2003) and the Behavior Rating Inventory of Executive Function, Second Edition, (BRIEF-2) (Hendrickson & McCrimmon, 2019).

The WISC-V is a comprehensive measure of overall intellectual ability, and five specific domains (verbal comprehension, visual spatial, fluid reasoning, working memory, processing speed). The BASC-3 is a questionnaire assessment of adaptive and maladaptive behaviour and emotions, including mood (e.g. depression and anxiety) in youth consistent with DSM-5 classifications. The BRIEF-2 assesses executive functioning skills in home and school environments. All assessments were supervised by a trained clinical health psychologist.

### Statistical analysis

Non-parametric Kruskal-Wallis test was performed to test for group differences in age, sex and BMI between PWS and the controls. Further statistical analyses were performed using MATLAB version R2022b, JASP version 0.18.0 (JASP, 2023), and the Network-Based Statistic (NBS) toolbox (Zalesky et al., 2010). False positives due to multiple comparisons were controlled using the Benjamini-Hochberg false discovery rate (FDR-BH) at a significance threshold of *p* < 0.05. Analysis of variance (ANOVA) comparisons were used to test for group differences between PWS and the controls on standardized scores from the WISC-V, , BASC-3, and BRIEF-2.

### MEG acquisition and source estimation

Five minutes of eyes open *Inscapes* “pseudo-resting-state” resting state MEG data were acquired on a 151 channel CTF system at the Hospital for Sick Children in Toronto. *Inscapes* is similar to a naturalistic viewing paradigm (Vanderwal et al., 2015), and is a video of moving computer generated images with an accompanying audio soundscape. Participants were instructed to “remain awake and in a state of rest, allowing their mind to wander.” *Inscapes* has been shown to recapitulate resting state network activity analogous to ‘traditional’ fixation cross resting state, and pertinent to the PWS group, is also known to increase participant compliance / tolerance with scan parameters and subsequently reduce the amount of head motion artefacts (Vandewouw et al., 2021). Participants were scanned in the supine position with fiducial coils attached at the nasion and bilateral preauricular points for continuous head motion recording. MEG data was sampled at 600 Hz, and continuous, sensor level data were filtered using a 4th-order Butterworth band-pass filter to retain frequencies between 1 and 150 Hz. Then, a Fourier transform notch filter was applied to eliminate noise at the power line harmonics of 60 and 120 Hz.

Time-series data were divided into 10 s epochs and the maximum number of epochs retained from the 5-minute recording based on the following exclusion criteria: (a) head position did deviate more than 105 mm during a given epoch; (b) epochs that contained SQUID jumps (or signal jumps) in the MEG signal that exceeded ±2 pT.

We employed Independent Component Analysis (ICA) using the ‘fastica’ algorithm in FieldTrip (Oostenveld et al., 2011) to extract and remove cardiac and eye movement artifacts. Following ICA decomposition, visual inspection of the components identified those corresponding to artifacts. These components were then manually removed by an experienced analyst. The resulting artifact-free data (post-ICA cleaned) was used for subsequent analyses.

To pinpoint the sources of activity in the cerebral cortex, we defined 90 cortical areas based on the Automated Anatomical Labelling Atlas (AAL) (Tzourio-Mazoyer et al., 2002). We then employed a linearly constrained minimum variance (LCMV) beamformer (Van Veen et al., 1997) to estimate the electrophysiological activity within these predefined regions of interest (ROIs) in the brain at the centroid of the parcel. MEG data co-registration involved creating a single-shell head model for each participant using age-appropriate anatomical T1-weighted templates (Evans & Brain Development Cooperative Group, 2006). This facilitated the generation of a forward solution required for the beamformer analysis. A common spatial filter was calculated for each seed location, and a covariance matrix was generated (regularized with a 5% Tikhonov factor) using the epoched, artifact-free sensor-level data. Finally, source-level broadband time series were computed for the centroid of each ROI (Sekihara et al., 2001).

### MEG signal analysis

Broadband-limited cortical oscillatory activity within each brain region was calculated based on the power spectral density (PSD) for the 78 cortical areas from the 90 AAL atlas, removing subcortical regions from any further analysis. Time series data for each region was z-score normalized (via mean centering and variance normalization), and the PSD for each epoch was calculated using Welch’s method and separated into canonical frequency bands: Delta (1-3 Hz), Theta (4-7 Hz), Alpha (8-14 Hz), Beta (15-30 Hz), Low Gamma1 (30-55 Hz), Low Gamma2 (65-80 Hz), and High Gamma (80-150 Hz) ranges. Finally, the power values for each frequency band were averaged across remaining epochs for subsequent statistical analysis.

For functional connectivity, the weighted phase lag index (wPLI) was employed (Vinck et al., 2011) to assess functional connectivity between brain regions. We used 76 of the AAL atlas nodes, including networks: the anterior DMN, aDMN (22 nodes); posterior DMN, pDMN (18 nodes); sensorimotor network, SMN (12 nodes); salience network, SN (10 nodes); memory network, MN (32 nodes); visual network, VN (12 nodes); central executive network, CEN (8 nodes); and attention network, AN (18 nodes). wPLI quantifies phase synchronization between brain areas by leveraging the magnitude of the cross-spectrum’s imaginary component (Lau et al., 2012; Vinck et al., 2011), providing a measure sensitive to neural ‘communication-through-coherence’. Calculations for wPLI were performed between all pairwise connections for the nodes included in the resting state networks, for each frequency band; with values ranging from 0 (indicating desynchronisation between regions i.e. low functional connectivity) to 1 (high synchronisation, consistent with high functional connectivity).

Graph measures of network topology were calculated using the Brain Connectivity Toolbox (Rubinov & Sporns, 2010) using the functional connectivity matrices to define all analysed brain networks used as inputs (e.g. 76x76 node matrices. Briefly, we examined measures of centrality with Eigenvector Centrality, Assortativity, and Hubness of the networks (Rubinov & Sporns, 2010). Eigenvector centrality, or eigencentrality, is a measure that calculates the relative prominence of nodes within a network. Eigencentrality differs from other centrality measures in that it extends beyond looking at direct connections of a node and incorporates the influence of a node’s neighbors’ connections as well. Hubness refers to a small number of nodes having a large number of connections in comparison to other nodes. In the context of neuroimaging, hubs are brain regions that have a high number of connections to other regions. Assortativity measures the tendency of nodes with similar properties to connect with each other. High assortativity within a network indicates that nodes with high degrees (hubs) tend to connect with other high-degree nodes (or hubs). Together, these measures could be applied to characterize the developing brain’s functional architecture by identifying differences in network formation and connection, as well as shedding light on organization of functional networks within the brain.

### MEG statistical analysis

ANOVAs were used to test for group differences between individuals with PWS and controls in power (i.e. oscillatory activity) across all seven frequency bands across the left and right hemispheres by calculating the average power for each AAL atlas node in the left and right frontal, temporal, parietal, and occipital lobes, and subcortical regions (grouping together the hippocampus, amygdala, caudate, pallidum, and thalamus nodes).

The Network Based Statistics toolbox (Zalesky et al., 2010) was used to test group differences in functional connectivity within eight functional networks, across the seven MEG frequency bands using wPLI as edge weights (statistical model: threshold = 1.8; t-test; 5000 permutations; NBS method; Extent component size).

### Brain function and outcome association analyses

As a post-hoc analysis, correlations were conducted between brain measures (MEG intranetwork functional connectivity and graph theory metrics) that showed significant group differences and neuropsychological outcomes (WISC-V, BASC-3, BRIEF-2).

## Results

### Poorer neuropsychological outcomes in children with PWS compared to controls

Children with PWS showed extensive cognitive impairment with significantly lower performance compared to the controls across all summary indices and subtests of the WISC-V, including the full-scale IQ score and verbal comprehension, visual spatial index, fluid reasoning, working memory, and processing speed indices (**Table 1**).

**Table 1:** Controls and PWS standardized WISC-V scores.

| Wechsler Intelligence Scale for Children<br>5<sup>th</sup> Edition (WISC-V): Standard scores | Mean ± standard deviation | | Group effects (all significant at FDR-BH $p < 0.05$) | | |
| --- | --- | --- | --- | --- | --- |
| | Controls | PWS | F | (p) | Effect size $\eta^2$ |
| **Wechsler Intelligence Scale for Children, 5<sup>th</sup> Edition (WISC-V): Standard scores** |  |  |  |  |  |
| *Full scale IQ* | 99 ± 18 (n = 14) | 47 ± 10 (n = 12) | 78.17 | **5.14E-9** | 0.77 |
| *Verbal comprehension index* | 102 ± 17 (n = 14) | 54 ± 11 (n = 12) | 69.81 | **1.45E-8** | 0.74 |
| Similarities | 10 ± 3.4 (n = 15) | 2.1 ± 2.1 (n = 13) | 51.45 | **1.29E-7** | 0.66 |
| Vocabulary | 11 ± 3.0 (n = 15) | 2.3 ± 1.7 (n = 12) | 87.32 | **1.24E-9** | 0.78 |
| *Visual spatial index* | 100 ± 16 (n = 14) | 57 ± 14 (n = 11) | 49.70 | **3.50E-7** | 0.68 |
| Block design | 10 ± 3.8 (n = 15) | 2.7 ± 2.4 (n = 13) | 39.73 | **1.13E-6** | 0.60 |
| Visual puzzles | 11 ± 2.2 (n = 15) | 2.9 ± 2.3 (n = 13) | 85.43 | **1.06E-9** | 0.77 |
| *Fluid reasoning index* | 98 ± 19 (n = 14) | 59 ± 11 (n = 13) | 40.54 | **1.39E-6** | 0.63 |
| Matrix reasoning | 10 ± 3.7 (n = 15) | 1.9 ± 1.4 (n = 13) | 54.96 | **7.15E-8** | 0.68 |
| Figure weights | 9.5 ± 3.2 (n = 15) | 3.8 ± 2.6 (n = 13) | 27.34 | **1.84E-5** | 0.51 |
| *Working memory index* | 95 ± 12 (n = 14) | 56 ± 9.2 (n = 13) | 85.26 | **1.56E-9** | 0.77 |
| Digit span | 9.4 ± 2.4 (n = 15) | 1.5 ± 0.97 (n = 13) | 125.63 | **1.87E-11** | 0.83 |
| Picture span | 9.3 ± 2.3 (n = 15) | 3.7 ± 2.3 (n = 13) | 41.72 | **7.61E-7** | 0.62 |
| *Processing speed index* | 99 ± 13 (n = 14) | 54 ± 11 (n = 13) | 90.13 | **9.05E-10** | 0.78 |
| Coding | 9.3 ± 2.6 (n = 15) | 1.7 ± 1.5 (n = 13) | 86.78 | **9.08E-10** | 0.77 |
| Symbol search | 9.4 ± 2.8 (n = 14) | 3.1 ± 2.0 (n = 13) | 45.22 | **4.78E-7** | 0.64 |

On the parent-rated BASC-3, children with PWS were rated lower than controls on the summary indices of externalizing problems – specifically hyperactivity and conduct problems, but not in aggression or internalizing problems. BASC-3 scores were significantly lower in PWS children in the domains of behavioural symptoms, and adaptive skills (**Table 2**).

**Table 2:** Controls and PWS standardized parent-rated BASC-3 scores.

| Behavior Assessment System for Children 3rd Ed (BASC-3):<br>Parent-rated t scores | Mean ± standard deviation |  | Group effects (significant at FDR-BH p<0.05) |  |  |
| --- | --- | --- | --- | --- | --- |
| | Controls (n = 15) | PWS (n = 15) | F | (p) | Effect size $\eta^2$ |
| *Externalizing problems* | 49 ± 5.9 | 59 ± 11 | 10.85 | **0.003** | 0.28 |
| Hyperactivity | 48 ± 8.5 | 63 ± 14 | 13.02 | **0.001** | 0.32 |
| Aggression | 50 ± 5.7 | 57 ± 13 | 3.31 | ns | 0.11 |
| Conduct problems | 48 ± 6.1 | 56 ± 11 | 5.31 | **0.029** | 0.16 |
| *Internalizing problems* | 49 ± 7.3 | 56 ± 14 | 3.80 | ns | 0.12 |
| Anxiety | 50 ± 8.3 | 56 ± 13 | 2.46 | ns | 0.08 |
| Depression | 49 ± 6.4 | 55 ± 11 | 3.80 | ns | 0.12 |
| Somatization | 49 ± 8.0 | 56 ± 15 | 2.72 | ns | 0.09 |
| *Behavioural symptom index* | 48 ± 5.9 | 64 ± 12 | 23.92 | **3.74E-5** | 0.46 |
| Atypicality | 46 ± 5.3 | 67 ± 16 | 23.81 | **3.86E-5** | 0.46 |
| Withdrawal | 49 ± 8.8 | 63 ± 15 | 8.65 | **0.006** | 0.24 |
| Attention problems | 46 ± 8.4 | 62 ± 11 | 20.67 | **9.58E-5** | 0.43 |
| *Adaptive skills* | 57 ± 6.7 | 37 ± 7.3 | 57.88 | **2.75E-8** | 0.67 |
| Adaptability | 57 ± 7.3 | 41 ± 9.0 | 25.68 | **2.30E-5** | 0.48 |
| Social skills | 57 ± 7.0 | 41 ± 8.1 | 33.40 | **3.21E-6** | 0.54 |
| Leadership | 56 ± 8.2 | 37 ± 7.3 | 44.47 | **3.09E-7** | 0.61 |
| Activities of daily living | 54 ± 6.8 | 38 ± 8.4 | 30.84 | **6.13E-6** | 0.52 |
| Functional communication | 55 ± 6.7 | 35 ± 8.0 | 51.89 | **7.70E-8** | 0.65 |

Children with PWS also showed widespread executive control dysfunction as demonstrated by significantly lower parent-ratings relative to controls on the global composite score and across all summary indices including behaviour, emotional, and cognitive regulation (**Table 3**).

**Table 3:** Controls and PWS standardized parent-rated BRIEF-2 scores.

| Behavior Rating Inventory of Executive Function 2<sup>nd</sup> (BRIEF-2): Parent-rated standard scores | Mean ± standard deviation | | Group effects (all significant at FDR-BH $p < 0.05$) | | |
| --- | --- | --- | --- | --- | --- |
| | Controls (n = 15) | PWS (n = 15) | F | (p) | Effect size $\eta^2$ |
| *Global executive composite* | 48 ± 6.7 (n = 14) | 69 ± 11 (n = 15) | 30.90 | **7.75E-6** | 0.54 |
| *Behaviour regulation index* | 46 ± 5.8 (n = 15) | 67 ± 12 (n = 15) | 39.69 | **8.19E-7** | 0.59 |
| Inhibition problems | 46 ± 5.9 (n = 15) | 65 ± 12 (n = 15) | 32.73 | **3.89E-5** | 0.54 |
| Self-monitor problems | 47 ± 6.9 (n = 15) | 67 ± 12 (n = 15) | 31.90 | **4.74E-6** | 0.53 |
| *Emotional regulation index* | 48 ± 8.2 (n = 14) | 72 ± 14 (n = 15) | 31.61 | **5.75E-6** | 0.54 |
| Shifting problems | 46 ± 7.4 (n = 14) | 74 ± 15 (n = 15) | 38.57 | **1.22E-6** | 0.59 |
| Emotional control problems | 50 ± 9.6 (n = 15) | 67 ± 12 (n = 15) | 19.22 | **1.49E-4** | 0.41 |
| *Cognitive regulation index* | 49 ± 7.2 (n = 14) | 63 ± 12 (n = 14) | 14.92 | **6.68E-4** | 0.37 |
| Task-completion/-monitor | 49 ± 7.4 (n = 15) | 63 ± 8.5 (n = 15) | 25.40 | **2.49E-5** | 0.48 |
| Working memory | 49 ± 7.4 (n = 15) | 64 ± 12 (n = 15) | 16.68 | **3.36E-4** | 0.37 |
| Planning/organization problems | 47 ± 7.4 (n = 15) | 60 ± 14 (n = 15) | 10.64 | **0.003** | 0.28 |
| Initiate problems | 50 ± 8.5 (n = 15) | 64 ± 11 (n = 15) | 13.16 | **0.001** | 0.32 |
| Organization of materials problems | 49 ± 5.7 (n = 15) | 57 ± 11 (n = 14) | 5.78 | **0.023** | 0.18 |

### No regional differences in cortical oscillatory activity between PWS and controls

ANOVAs revealed no significant group differences between PWS and controls for lobe-wise and subcortical oscillatory activity across the seven frequency bands tested.

### Intranetwork dysconnectivity in visual, attention, and default mode networks, and altered network structure in PWS

MEG functional connectivity using wPLI revealed significantly lower neural synchrony in the PWS group relative to the control group in the Visual, pDMN and attentional networks (p < 0.05, FDR-corrected). The PWS group showed lower connectivity than controls in the beta range in the visual network (15-25 Hz; 8 nodes and 10 edges; **Figure 1a**), theta in the posterior default mode network (3-7 Hz; 13 nodes and 15 edges; **Figure 1b**), and in the low gamma range in the attentional network (65-80 Hz; 17 nodes and 21 edges; **Figure 1c**).

**Figure 1.**
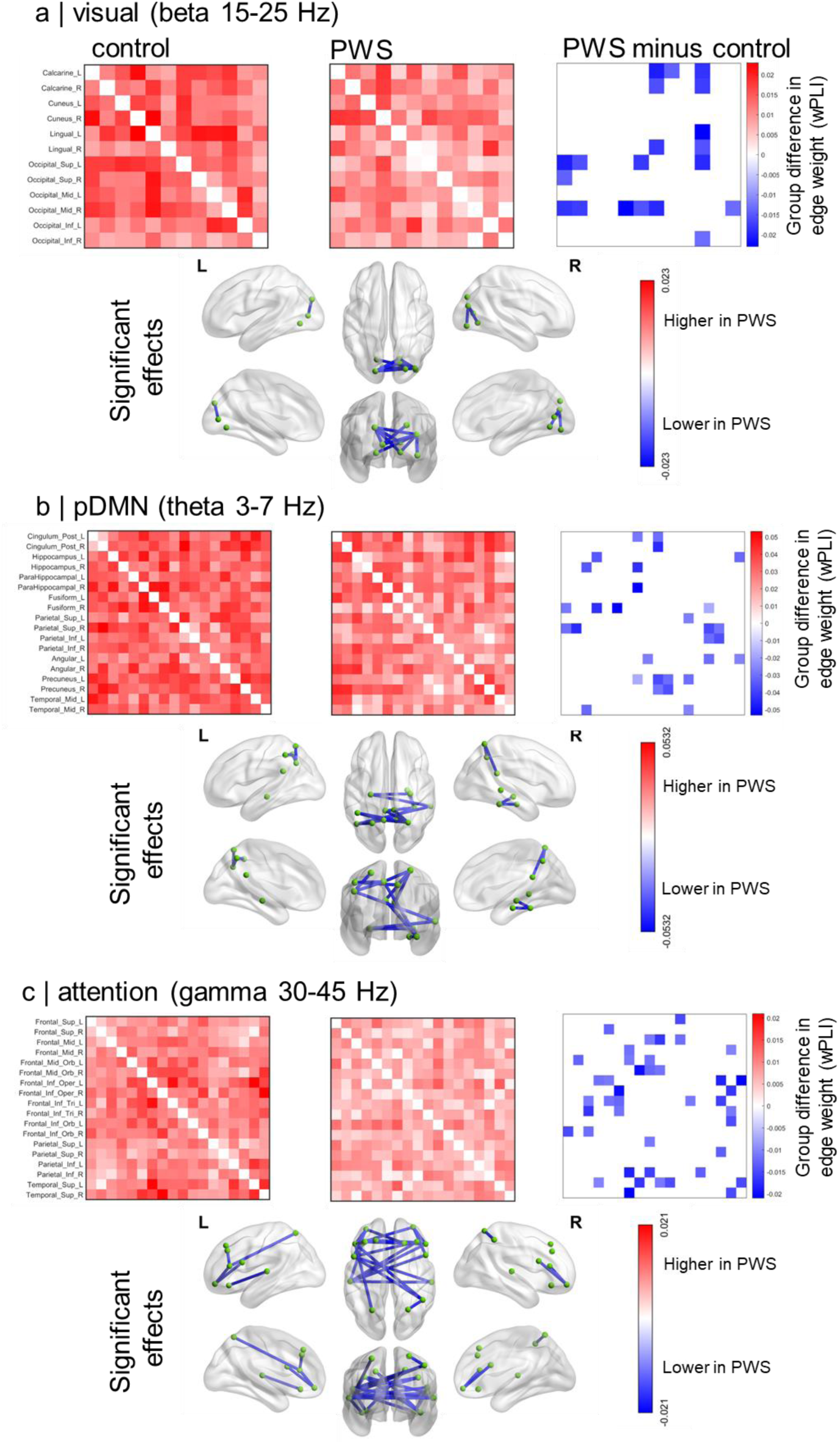
PWS is associated with reduced neural connectivity in the visual, posterior default mode, and attention networks. MEG wPLI dysconnectivity was evident in PWS in the **(a)** visual network mediated by beta (15-25 Hz) synchrony**, (b)** pDMN desynchrony in the theta range (3-7 Hz), and **(c)** in the attention network in the gamma range (30-45 Hz).

The PWS group also showed altered network organisation - using the entire functional connectome that made up the 8 pre-defined intrinsic brain networks - where average hubness mediated by low-frequency synchronisation in the delta range (1-3 Hz) was higher in PWS (**Figure 2a**; *p* = 0.02), and eigencentrality was lower in the PWS group (**Figure 2b**; *p* = 0.03).

**Figure 2.**
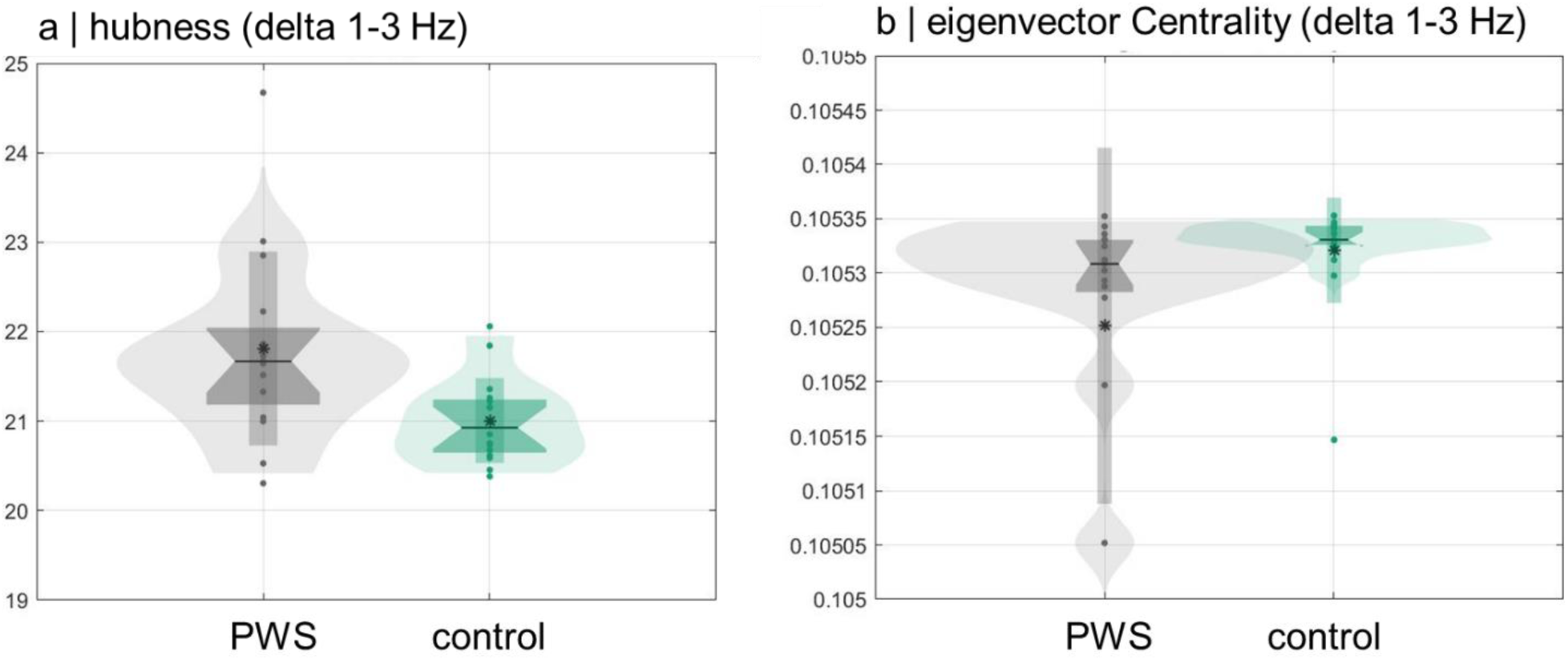
PWS is associated with altered network topological structure. Delta (1-3 Hz) mediated neural synchrony revealed increased hubness (a) in PWS and decreased eigencentrality (b).

### Correlation of network functional connectivity and graph measures with neuropsychological outcomes

Finally, we examined the relationship between MEG measures that showed significant group differences (visual, attentional, pDMN connectivity, and hubness and eigencentrality) across neuropsychological outcomes (**Figure 3**). Despite some moderate effect sizes (e.g. r coefficients), none of the correlations survived multiple comparison corrections.

**Figure 3.**
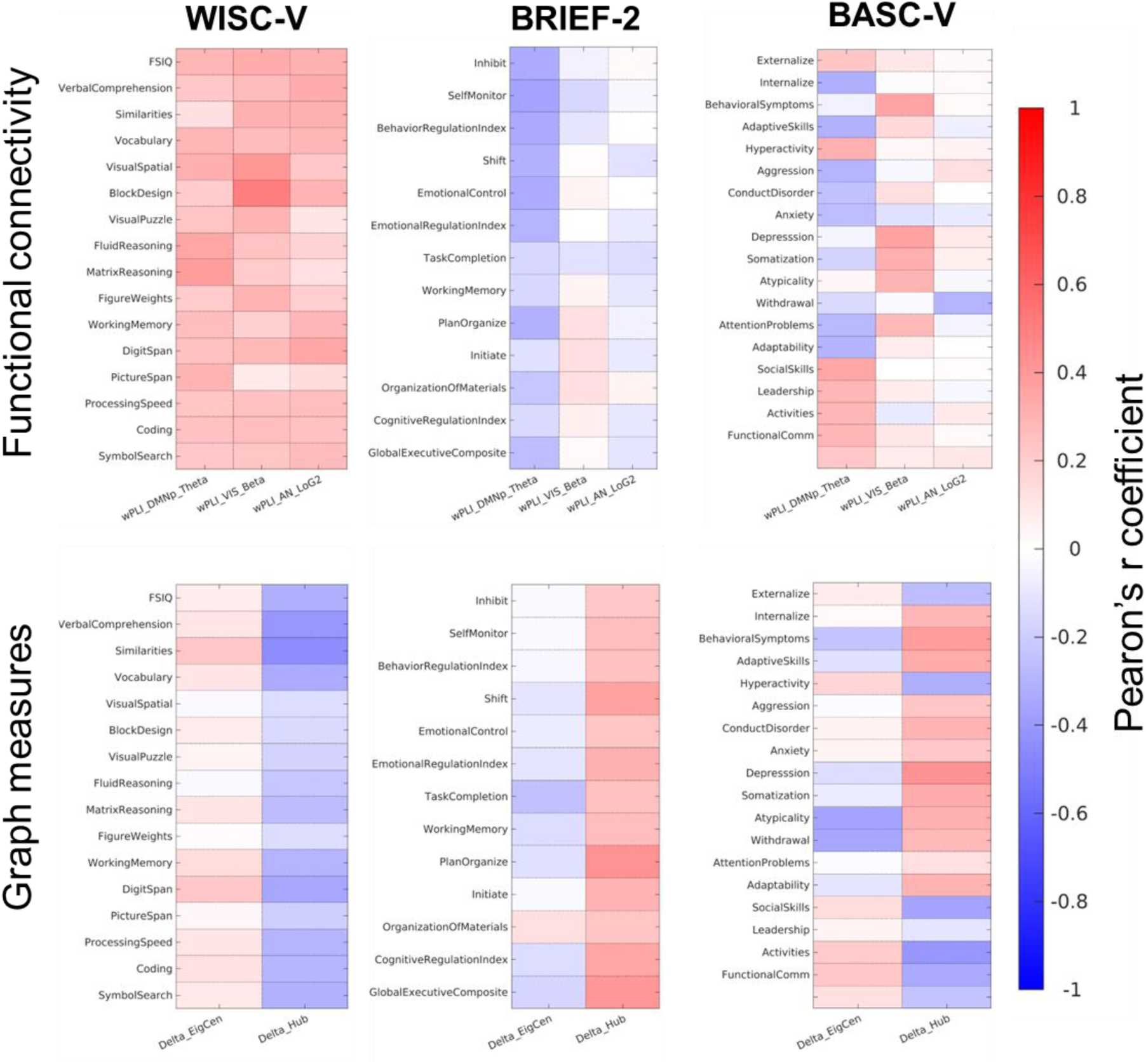
MEG network and graph structure correlations with neuropsychological outcomes. Despite several moderate correlations between functional connectivity/network structure and outcome measures, none of the measures survived multiple comparison correction.

## Discussion

### Summary

We present the first study of neuropsychological outcomes across a broad range of cognitive, emotional, and behavioural measures and neurophysiological activity and functional connectivity in a developmental cohort of Prader Willi Syndrome, against a BMI-controlled comparator group. As expected, we found large effects in terms of cognitive functioning, with reduced general cognitive abilities and significant behavioural issues in the PWS cohort, and also observed decreased functional connectivity, mediated by neural synchrony in the visual, attentional and posterior default mode networks. Further, MEG revealed changes in the topology of brain networks - but there were no statistically significant relations between neuropsychological outcomes and the MEG measures tested.

### Neuropsychological dysregulation in PWS

Through a neuropsychology assessment using the WISC-V, BASC-3, and BRIEF-2, we assessed cognitive, emotional (e.g. mood, stress), and behavioural domains.

### Cognition

Our findings reveal significant cognitive impairment in youth with PWS. On the WISC-V, the PWS cohort rated lower on full-scale IQ (FSIQ) and all cognitive indices (verbal comprehension, visual spatial, fluid reasoning, working memory, processing speed) relative to controls. This is in keeping with previous research; Whittington et al. similarly found that individuals with PWS scored lower on all subscales of their age appropriate Weschler intelligence test and had a downward shift in the normal distribution of FSIQ scores by approximately 40 points (Whittington et al., 2004). Although global functioning is impaired, it is important to consider that ability can vary between individuals depending on environmental or other genetic factors (Whittington & Holland, 2017).

### Emotions and Behaviour

The parent-reported BASC-3 scores indicated that children with PWS display significantly more externalizing behaviours compared to their peers; no concerns with internalizing symptoms (e.g., mood, anxiety) were endorsed by parents. This is discordant from a previous study by Skokauskas et al. (2012) that showed no significant difference in externalizing problems between pediatric PWS and control participants, but higher internalizing problems in PWS on the parent-report Child Behaviour Checklist 6-18 (CBCL/6-18). Although the BASC-3 and CBCL are distinct assessment instruments, internalizing and externalizing scores were shown to be highly correlated between the two in a sample of preschoolers born preterm, which may not be comparable to our population (Camerota et al., 2024). Our findings may be unexpected given the anxious tendencies seen in PWS, however our sample sizes were small, and participants recruited were well supported by mental health resources.

Participants with PWS scored highest on Atypicality and Withdrawal on the BASC-3, similarly to a report of children and adolescents with autism spectrum disorder (ASD) (Zhou et al., 2022). This provides further support for the behavioural overlap between ASD and PWS (Bennett et al., 2015). Participants with PWS were also ranked lower on the Adaptive Skills Composite and all subscales (adaptability, functional communication, leadership, social skills, activities of daily living), indicating challenges with adapting readily to changes in environment. This is consistent with task, routine, and thought rigidity seen in PWS (Schwartz et al., 2021), and in children with global developmental delay or severe intellectual disability more broadly (Moeschler et al., 2014). Future studies comparing these populations would delineate whether there are specific difficulties in PWS profiles, or if our findings are a reflection of global developmental delay.

### Executive Functioning in Daily Life

To our knowledge, we are the first group to use the BRIEF-2 in a study of PWS, which assesses executive functioning in home environments. The PWS cohort exhibited widespread executive control dysfunction as demonstrated by significantly lower parent-ratings relative to controls on the global composite score and across all summary indices including behaviour, emotional, and cognitive regulation. Those with PWS were rated most negatively on the Shift scale, part of the Emotional Regulation Index, which aligns with previous research highlighting task-switching as a particular area of executive dysfunction in PWS (Woodcock, Oliver, et al., 2009). Deficits in shifting ability can manifest as reduced efficiency or stagnation when problem-solving, perseverative behaviours, and marked resistance to change, all which have been reported in individuals with PWS.

### Reduced executive functioning and altered brain activity in PWS

It is well-established that individuals with PWS present with executive dysfunction and differences in behaviour. Indeed, our results demonstrated that our group of children with PWS experience challenges with executive functioning skills in daily life as observed by their parents. Prior research has also shown that functional brain abnormalities are prominent in PWS and may underlie the cognitive and behavioral profile of the disorder. Abnormalities include hypothalamus and interconnected brain circuits, including those involved in appetite regulation, motivation and reward, decision making, planning and emotional areas, such as the prefrontal cortex, amygdala, insula, and nucleus accumbens (Zhang et al., 2013).

For the MEG data, our study found decreased functional connectivity in the visual, attention and pDMN, mediated by neural synchronisation amongst brain regions. Neural synchronisation is the mechanism by which distributed brain areas coordinate activity and communicate – ‘communication through coherence’ (Fries, 2005, 2015). Rhythmic activity in cortical microcircuits waxes and wanes in terms of membrane excitability, open and closing windows of opportunity for signals to be transmitted or integrated, allowing functionally disparate regions to align activity and form functional bran networks that underlie cognition and behaviour (Uhlhaas et al., 2009), including executive functions that emerge from theta phase synchrony between fronto-parietal regions (Mizuhara & Yamaguchi, 2007) and information encoding via theta in default mode regions (Johnson et al., 2023). In line with our results showing reduced theta synchrony in the DMN, other studies postulate that theta synchrony is a crucial mechanism that supports filtering of task-relevant and irrelevant information, particularly in the context of memory encoding and retrieval (Solomon et al., 2017). This may underlie the executive function deficits presented in people with PWS.

We also observed reduced high frequency gamma activity in PWS, particularly amongst frontotemporal brain areas that form part of the attention networks. High frequency gamma activity is thought generated by the interaction between excitatory pyramidal neurons and inhibitory interneurons (Buzsáki & Wang, 2012), acting as an integrative mechanisms for the focusing of attention (Doesburg et al., 2008) and the emergent of perceptual states (Uhlhaas et al., 2011). These reductions in gamma might reflects reductions in the integrative neurobiological processes required for coherent percepts and the orienting of attentional processes that broadly speak to neuropsychological deficits present in this group. In particular, impaired attentional mechanisms as a result of reduced gamma oscillations in frontotemporal regions suggest compromised ability to orient and maintain attentional focus in individuals with PWS (Schwartz et al., 2021). This supports previous fMRI studies that showed alterations in selective attention and cognitive control in people with PWS, particularly when faced with palatable food cues and environmental distractions (Holsen et al., 2006).

Moreover, we observed reduced beta synchronisation in visual areas in PWS. Beta synchrony in sensorimotor regions is proposed as a mechanism for the rhythmic top-down coordination of oculomotor signals feedback from frontal and cingulate regions, including the frontal eye fields and anterior cingulate cortex (Babapoor-Farrokhran et al., 2017), as well as integrating top down and bottom up sensory information (Richter et al., 2018). Ocular and perceptual deficits are well known in PWS, including reduced visual acuity, strabismus, and refraction (Hered et al., 1988), with dorsal visual stream deficits (the ‘where’ pathway) being particularly pronounced (Woodcock, Humphreys, et al., 2009).

### Limitations and Future Directions

Our study has a number of limitations. First, we present MEG data in the resting state rather than task-based (e.g. food cues, cognitive task paradigms). While measuring spontaneous functioning can reveal much about intrinsic neural activity, it fails to capture the full repertoire of cognitive and behavioural states. Nevertheless, the task-free design of resting state means that there are no explicit task instructions to follow and those individuals who might otherwise find tasks difficult can more easily comply with assessment. In this particular case, we found that the *Inscapes* paradigm was successful in capturing attention and minimising head motion artefacts in the PWS cohort. Nevertheless, correlating outcome measures with resting intrinsic brain function is beset with difficulties, and after correcting for multiple comparisons, we did not observe any significant effects (despite moderate effect sizes/degree of association). Future studies would employ multivariate approaches (such as partial least squares, PLS) that model latent variables and are ideal for working high dimensional data such as this.

Despite these limitations, our study presents a novel use of MEG in PWS, which proved to be an effective method of recording functional brain connectivity in this group that can sometimes be poorly compliant with scan acquisitions, and was here, well tolerated by individuals.

Leveraging MEG may be a promising alternative for future brain studies in children with behavioural challenges given that it is less invasive than fMRI and therefore may lend itself well to administering task-based paradigms within it. Future neurophysiological research in this population should also consider the genetic subtypes of PWS (paternal chromosome 15 deletion versus uniparental disomy) as recent literature has shown phenotypic differences between the subtypes (Butler et al., 2004).

## Conclusions

This study reports neuropsychological dysregulation, as well as reduced functional connectivity via neural synchronisation in a developmental cohort of youth with Prader Willi Syndrome, against a BMI-controlled comparator group. We observed neural deficits in key brain networks involved in attentional control, executive functioning, and visual perceptual processes, suggesting that an inability to effectively coordinate neural synchrony in core brain networks, at least in part, underlies some of the cognitive, behavioral, and emotional issues exhibited by individuals with PWS.

## Data Availability

All data produced in the present study are available upon reasonable request to the authors

## Acknowledgements

We would like to thank study participants and their caregivers for their participation in the study.

